# Evidence needs and community involvement in policy decisions for vaccine-preventable diarrhoeal infections among children under the age of five years: Stakeholder engagement in Ethiopia, Kenya, and Malawi

**DOI:** 10.1101/2025.05.27.25328430

**Authors:** Chikondi Mwendera, Mackwellings Phiri, Rahma Osman, Shewit Weldegebriel, Beatrice Ongadi, Catherine Beavis, Jessica A. Fleming, Dan Hungerford, Nigel Cunliffe, Deborah Nyirenda, Neil French, GHRG-GI Consortium

**Author notes:** Joint Senior authors (Deborah Nyirenda, Neil French). Members of the GHRG-GI Consortium listed at the end of the paper.

## Abstract

Diarrhoeal diseases are a leading cause of morbidity and mortality among children under five years in low- and middle-income countries (LMICs) with *Shigella* and Enterotoxigenic *Escherichia coli* (ETEC), both targets of ongoing vaccine development, playing major roles. To inform the introduction of vaccines against these pathogens in Ethiopia, Kenya, and Malawi, we assessed evidence needs for technical policy decisions and community engagement in national policy development processes. Using in-depth interviews (IDIs) with 27 institutional stakeholders (health ministries, researchers, and partners) and focus group discussions (FGDs) with community representatives (one per country), we collected data from May 2023 to May 2024 and undertook thematic analysis.

Institutional stakeholders emphasised the need for robust, localised evidence on disease burden, vaccine efficacy, safety, cost-effectiveness, programmatic feasibility, alongside structured mechanisms to integrate evidence into policy. They noted the absence of diarrhoeal disease-specific technical working groups in their countries and advocated for their establishment. Meanwhile, community representatives expressed vaccine acceptance but stressed the importance of clear communication of information delivered by trusted health workers and leaders. They highlighted the need for comprehensive information on new vaccines, particularly regarding their safety and potential side effects. While community involvement in national policy-making processes was deemed low, participants highlighted the value of meaningful engagement to ensure ownership and uptake.

The findings highlight the importance of robust evidence generation, effective communication, and inclusive community engagement to facilitate the successful introduction of *Shigella* and ETEC vaccines. Addressing these gaps can accelerate adoption, improve readiness, and foster trust in vaccination programmes.

## Introduction

Diarrhoeal diseases cause approximately 500,000 annual deaths among children under five years, with the heaviest burden falling on low- and middle-income countries (LMICs), particularly in sub-Saharan Africa (SSA) (1, 2). In SSA, rotavirus, *Shigella*, and Enterotoxigenic *Escherichia coli* (ETEC) are leading pathogens, fuelled by persistent gaps in water, sanitation, and hygiene (WASH) infrastructure (2–4). While oral rehydration therapy and intravenous fluids for severe cases of diarrhoea, and antibiotics for *Shigella* and ETEC remain key treatments, their effectiveness is limited by poor access and antimicrobial resistance (AMR) (5–8). Vaccines offer critical protection, as demonstrated by the success of rotavirus vaccine (RVV) programmes in Malawi, Ethiopia, and Kenya, which reduced rotavirus-associated hospitalisations, following their introduction in 2012, 2013, and 2014 respectively after the 2009 World Health Organization (WHO) recommendation (9–14). However, modest RVV effectiveness in LMICs and uneven vaccine coverage persist, leaving rotavirus a leading cause of childhood diarrhoea (15, 16).

Beyond rotavirus, *Shigella* and ETEC are major contributors to childhood diarrhoeal diseases in LMICs (17, 18). The rising burden of these pathogens, compounded by AMR, has prompted the WHO to prioritise the development of vaccines against them (19, 20). However, timely vaccine introduction is critical. For instance, since the WHO recommendation, the RVV introduction took three, four, and five years in Malawi, Ethiopia, and Kenya, respectively (10, 11, 14, 21). Such delays, more common in LMICs than high-income countries (22), represent missed opportunities for community health benefits and hinders progress towards global targets, including the immunisation-related UN Sustainable Development Goals (SDGs) (23). Addressing these delays requires governments to ensure that decisions on vaccine introduction are impartial, timely and evidence-based (24, 25).

Community engagement is proven to strengthen health interventions, particularly during implementation (26, 27). However, meaningful participation in national policy-making processes in Africa is often limited by token consultations, power imbalances, and weak decentralisation (28, 29). Bridging these gaps requires integrating communities into policy design (not just during implementation), to warrant sustainable and equitable health outcomes.

To ensure readiness for future *Shigella* and ETEC vaccines, LMICs must strengthen preparation, while improving RVV coverage. As part of the NIHR Global Health Research Group on Gastrointestinal Infections (GHRG-GIs), we conducted in-depth interviews (IDIs) with institutional stakeholders (ISs) in Ethiopia, Kenya, and Malawi to assess evidence needs for *Shigella* and ETEC vaccine introduction. This work aligns with the group’s broader efforts to generate evidence, build capacity, and engage communities for childhood enteric vaccines in eastern and southern Africa. Additionally, focus group discussions (FGDs) with community engagement groups gathered local perspectives on community participation in vaccine policy processes and strategies to foster community ownership and acceptance.

## Methods

### Study design

This qualitative study employed a phenomenological approach (30) to explore participants’ experiences in policy processes.

#### Participants, setting, and recruitment

##### Institutional stakeholders (ISs)

We purposively sampled ISs from Ethiopia, Kenya, and Malawi with expertise in childhood diarrhoeal vaccine policies and implementation, supplemented by snowball sampling. Participants included representatives from ministries of health (MoH), WHO country offices, national public health and technical advisory bodies, immunisation programme managers, researchers, and clinicians. Selection was based on institutional knowledge, decision-making roles, experience, and relevant expertise.

##### CEI groups

The GHRG-GI prioritises community engagement and involvement (CEI) by fostering collaboration between researchers and local communities in high-density urban areas with limited infrastructure and high infectious disease burdens. In Ethiopia, the study was conducted at Tikur Anbessa Specialized Teaching Hospital in Addis Ababa, focusing on Teklehaimanot and Kirkos sub-cities. Rapid urbanization here has led to overcrowding, inadequate clean water access, and high rates of infection (31). In Kenya, research was conducted at Mukuru Health Centre in Nairobi, targeting informal settlements (Kwa Reuben and Kwa Njenga). Poor water and sanitation drive diarrhoeal diseases (32). In Malawi the study was based at Bangwe Health Centre in Blantyre City’s informal settlements, where HIV/AIDS, malaria, tuberculosis, and diarrhoeal diseases are prevalent amid poor WASH conditions (33).

CEI group members (composed of community volunteers in each country) were selected through a participatory, context-sensitive process that emphasised community ownership and trusted representation. Communities nominated candidates through open dialogue, followed by a democratic selection based on criteria prioritising diversity, gender balance, and study relevance (e.g., caregivers, health workers, teachers). The process ensured inclusion of marginalised groups and those affected by gastrointestinal infections, resulting in a membership that reflected diverse social, cultural, and professional backgrounds.

#### Data collection

Data were collected between May 2023 and September 2024. The IDIs were conducted by the lead author (CM), a postdoctoral researcher in health policy analysis, using a structured guide with open-ended questions (Supplementary Appendix 1). The guide was reviewed by co-authors and piloted in each country and refined based on feedback. Interviews lasted up to 60 minutes and were conducted in English either in-person or virtually. The FGDs were facilitated by trained social scientists (SW, RO, and MP) in Ethiopia, Kenya, and Malawi, respectively. A topic guide (Supplementary Appendix 1) explored community awareness of childhood diarrhoeal disease, vaccine policies and perceived influence in decision-making. FGDs were conducted in local languages (Amharic in Ethiopia, Swahili in Kenya, and Chichewa in Malawi) and lasted approximately 90 minutes.

Discussions were audio-recorded and observational notes taken to contextualise the data, following participant briefing and informed consent. Ethical approval was obtained from review boards in each country: Addis Ababa University’s Department of Pediatrics (030-23-ped) in Ethiopia, KEMRI’s Scientific Ethics Review Unit (KEMRI/SERU/CMR/P00221-010-2022/4637) in Kenya, and Kamuzu University of Health Sciences (P.10/22/3790) in Malawi, as well as the University of Liverpool, UK (Ref: 12443).

#### Data analysis

Audio recordings from FGDs were transcribed verbatim and translated into English, while IDIs were transcribed verbatim. The authors involved in data collection conducted quality checks to ensure transcript accuracy and integrity. Qualitative analysis was performed thematically in NVivo 12 (34), using both inductive and deductive approaches guided by Braun and Clarke’s six-step framework (35). Two authors (CM and MP) independently coded the FGD and IDI transcripts to developed initial coding frameworks, which were then applied consistently across all transcripts. Codes were organised into categories, refined into subthemes, and synthesised into overarching themes for each dataset. Cros-country analysis identified shared perspectives through comparative synthesis of FGD and IDI data.

## Results

Twenty-seven IDIs were conducted with eight ISs each from Ethiopia and Kenya, and eleven from Malawi. Participants included immunisation program managers, clinicians, researchers, and members of technical working groups such as National Immunisation Technical Advisory Groups (NITAGs). Seven (25%) were female. All had over five years of relevant experience, and 52% (14/27) had more than 15 years (Table 1).

**Table 1:** In-depth interview stakeholder roles, gender, and experience, by country.

| Role, other capacity, and institution | Gender | Experience |
| --- | --- | --- |
| **Ethiopia** |  |  |
| Programme manager and NITAG member - MoH | Male | >12 years |
| Programme manager - NGO | Male | >5 years |
| Programme manager – UN Organisation | Male | >20 years |
| Programme manager – UN Organisation | Female | >11 years |
| Clinician and researcher – Academic Institution | Male | >15 years |
| Researcher and NITAG member – Academic Institution | Male | >15 years |
| Clinician and Researcher – Academic Institution | Male | >20 years |
| Programme manager and researcher - NGO | Male | >20 years |
| **Kenya** |  |  |
| Programme manager - MoH | Female | >15 years |
| Programme manager - NGO | Male | >11 years |
| Programme manager – UN Organisation | Male | >10 years |
| Programme manager – UN Organisation | Male | >10 years |
| Researcher and NITAG member – Academic Institution | Male | >20 years |
| Physician, researcher, and NITAG member – Academic Institution | Female | >10 years |
| Clinician and researcher – Academic Institution | Male | >15 years |
| Researcher – Research Institution | Male | >20 years |
| **Malawi** |  |  |
| Programme manager and NITAG member - MoH | Female | >5 years |
| Programme manager - MoH | Male | >15 years |
| Programme manager - NGO | Female | >10 years |
| Programme manager – NGO | Male | >10 years |
| Programme manager - UN Organisation | Female | >20 years |
| Programme manager – UN Organisation | Male | >15 years |
| Pediatrician and NITAG member – Academic Institution | Male | >20 years |
| Researcher – Academic Institution | Male | >15 years |
| Research and NITAG member – Research Institution | Male | >10 years |
| Researcher and NITAG member – Research Institution | Male | >10 years |
| Programme manager – Research Institution | Female | >5 years |

MoH Ministry of Health; NGO: Non-Governmental Organisations; NITAG: National Immunisation Technical Advisory Group; UN: United Nations

We conducted one FGD with the CEI groups per country. The FGDs included ten participants in Ethiopia (five females and five males), ten participants in Kenya (six females and four males), and seven participants in Malawi (four females and three males).

### Emerging themes

Four main themes related to evidence needs for technical decision-making and community engagement in national policy development processes emerged from the IDIs and the FGDs, including (1) policy involvement and evidence-related challenges, (2) evidence needs for policy decisions, (3) trusted sources of evidence, and (4) strategies for increasing evidence use. Sub-themes for each of these main themes are outlined in Figure 1.

**Figure 1:**
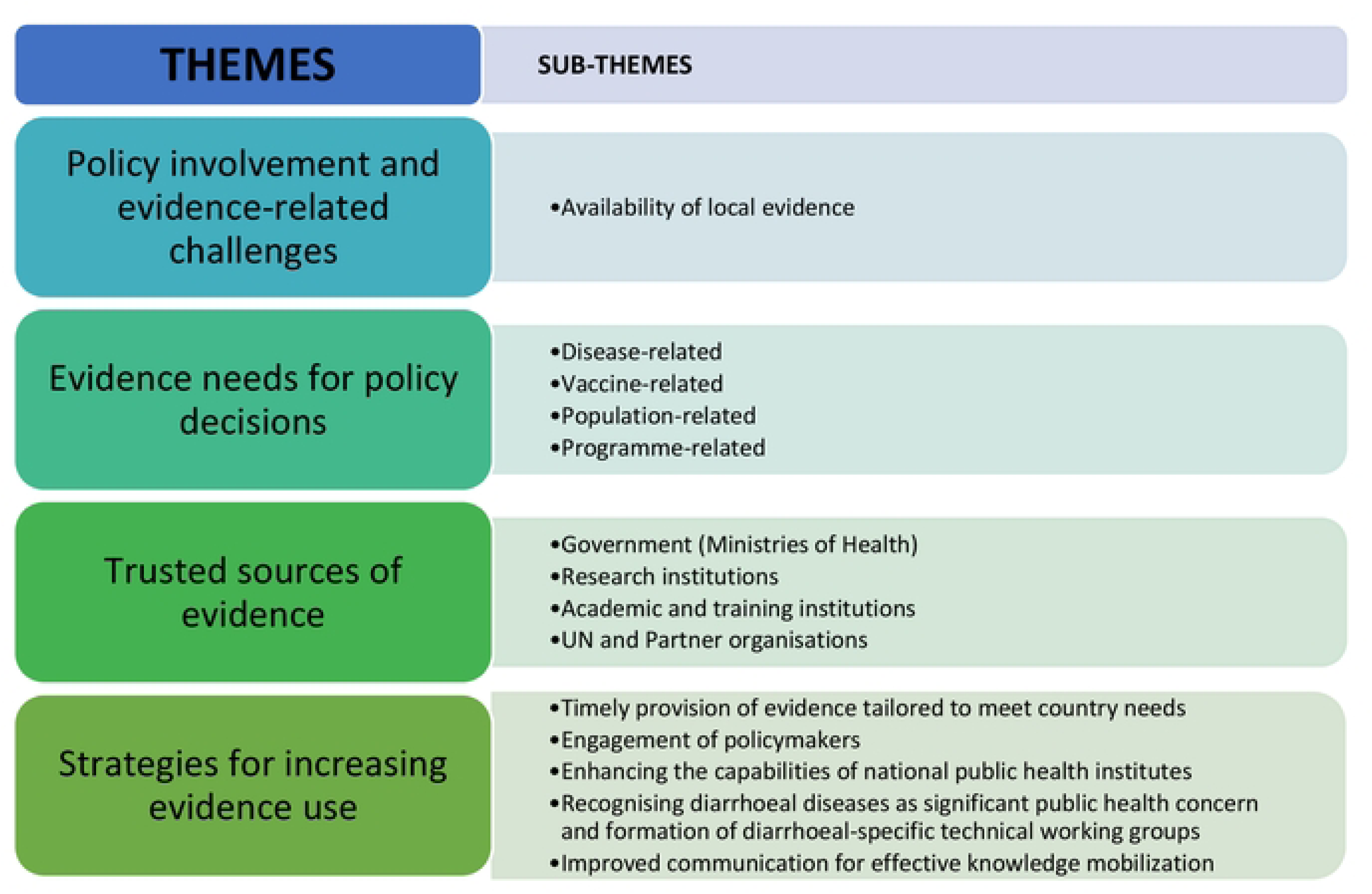
**Emerging themes and sub-themes on evidence needs for technical decision-making and community engagement in national policy development processes**.

#### 1. Policy involvement

ISs acknowledged their direct or indirect involvement in policy development processes for childhood vaccines in their respective countries. Their roles ranged from active participation in technical working groups (TWGs) such as NITAGs to drafting policy documents, providing consultations, and conducting research to inform policy decisions. This was expressed as follows:

> *“I sit in the Kenya National Immunisation Technical Advisory Group (KENITAG), which advises the ministry of health on all matters related to vaccines, including modifying existing schedules or introduction new vaccines”.* (**Institutional Stakeholder in Kenya)**

Similarly, an Ethiopian stakeholder highlighted their role in disease surveillance and influencing policy decisions:

> “*My centre was a diarrhoeal disease surveillance centre, so, we collected the surveillance data and based on that the burden of rotavirus as an important pathogen was confirmed. And that was strong evidence for the introduction of the rotavirus vaccine in this country”*. **(Institutional Stakeholder in Ethiopia)**

While in Malawi a stakeholder shared their experience in consulting for the MoH:

> “*I was a lead consultant in the ministry of health approval and national wide campaign for typhoid conjugate vaccine involving the preparation of all the necessary documentation, the campaign itself, engagement with relevant stakeholders, and training of the health workers and other relevant parties*”. **(Institutional Stakeholder in Malawi)**

ISs emphasised the necessity of local evidence for policy adoption, noting that while WHO guidelines offer direction, policymakers often require country-specific data before implementing recommendations. Without such evidence, regional data may be considered, but decisions are frequently postponed until national studies are available:

> “*Actually, having local evidence is one of the challenges most of the time when you introduce a vaccine…. we need to have some local evidence on how it will be effective in Ethiopia, given that Ethiopia has its own specific contexts*”. **(Institutional Stakeholder in Ethiopia)**

> “*You know, WHO can make a recommendation to say the vaccine can be rolled out. But it is not just an automatic thing; we still must see that there is a benefit of the intervention to the local communities. Otherwise, stakeholders can even shoot it down*”. **(Institutional Stakeholder in Malawi)**

While ISs participated in national policy development, community participants (CPs) across all three countries reported minimal engagement in national policy formulation, often associating policy participation with vaccine uptake. Although few had contributed directly to national policymaking, some supported policy implementation through local initiatives with health facilities, NGOs, and community organisations. For instance, a Malawian participant described joining community discussions on disease prevention:

> “*I have never been involved in any policy formulation process at national level, but I would say I took part at community level. I was called by the chief for our community to discuss about strategies for preventing diarrhoea and other water borne diseases in our community. I would say that I have attended such kind of meetings several times especially during the time when cholera was rampant here in Bangwe*”. **(FGD participant in Malawi)**

Similarly, a participant from Kenya highlighted their engagement in local initiatives:

> “*I’ve been involved in a few local initiatives that focus on specific health issues. Recently, I joined a community gardening project that aims to increase access to fresh produce and promote a healthier diet*”. **(FGD participant in Kenya)**

In Ethiopia, another participant noted their role in raising awareness on non-communicable diseases:

> “*I contributed to the development of some policies on educating people about risk factors of diabetes mellitus and hypertension in our community as well as how patients can take care of themselves*”. (**FGD participant in Ethiopia).**

Although rarely engaged at the national level, CPs strongly advocated for inclusion in policy development, believing this would enhance ownership and compliance. Many proposed indirect participation through pre-implementation studies exploring community perspectives before vaccine rollouts, viewing these as opportunities to inform policy and strengthen advocacy:

> “*In most cases before vaccines are administered to people, several studies are conducted for the government to decide how the vaccines should be administered …… Therefore, I would conclude that the outcomes of various studies on the way how vaccines should be administered define the ways through which a particular vaccine should be administered to people*”. **(FGD participant in Malawi)**

Similarly, a participant from Kenya emphasised the importance of advocacy strategies informed by community input:

> “*I believe it’s possible to influence the government’s decision by recommending community views through a comprehensive advocacy strategy*”. **(FGD participant in Kenya)**

Meanwhile in Ethiopia, participants suggested that direct involvement could be achieved if the government facilitated structured dialogues with community leaders or groups:

> “*To gain the entire community’s idea, it is preferable if they involve the leaders of the community. It will be fantastic to be involved in policy formulation through community leaders or with groups of mothers who have children under 5*”. **(FGD participant in Ethiopia)**

#### 2. Evidence needs for policy decisions

ISs identified four key evidence domains critical for informing policy decisions on new childhood enteric vaccines introduction into EPIs (Figure 2).

**Figure 2:**
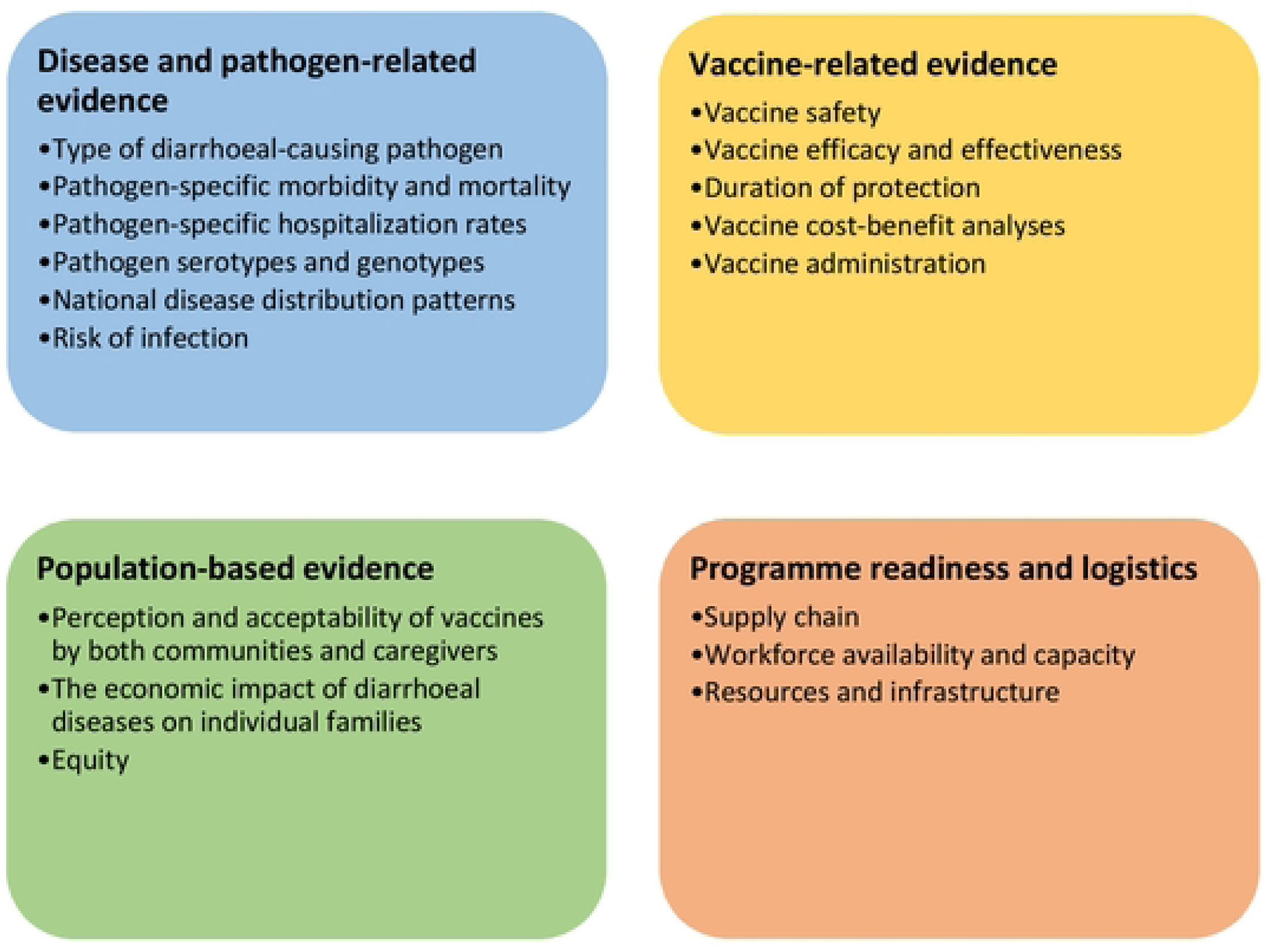
**Evidence domains to guide policy decisions.**

##### Disease and pathogen-related evidence

All ISs consistently highlighted that demonstrating significant disease burden is the most critical evidence needed to drive action. New vaccine policies depend on identifying urgent local health problems requiring intervention. For childhood diarrhoea, this requires precise pathogen identification, including serotypes and genotypes, along with clear epidemiological data on morbidity, hospitalisation, and mortality impacts:

> “*The main decision points are influenced by availability of data on the burden of disease. Basically, how many people get sick? How many of those also die? And you’ve got to compare it with other existing pathogens or other existing diseases*.” (**Institutional Stakeholder in Kenya**)

> “*First of all, I think a demonstration that there is burden…. So, I think that is a key data need, because if you don’t demonstrate burden, it becomes almost pointless to be working with vaccines and any other prevention activities because people don’t know that it is indeed a problem*”. **(Institutional Stakeholder from Malawi)**

> “*Epidemiological data is the best, because you must know the burden of illness. The morbidity and mortality data and the distribution of the disease are very important for deciding on a vaccine*.” **(Institutional Stakeholder from Ethiopia)**

##### Vaccine-related evidence

The ISs identified vaccine efficacy, effectiveness, and safety as key adoption drivers. They highlighted that WHO recommendations reinforce confidence in these standards, ensuring significant recipient benefits:

> “*You know, information about the vaccine and disease is very important, for example, from the vaccine side, safety, effectiveness, and efficacy are quite important. So, we must be able to convince the regulatory bodies, you know, about safety and efficacy. And we get comfort when WHO approves*.” **(Institutional Stakeholder in Ethiopia)**

Kenyan ISs highlighted vaccine affordability as a key concern during their transition from GAVI support to self-financing, stressing the need for cost-benefit analyses to prioritise vaccines among competing health interventions:

> “*So, we look at alternative costs… what are the alternatives and then we also weigh if there is treatment, which one is cheaper? Is it vaccines or treatment? So that in the end you’re able to rationalise and see whether vaccines would be cheaper*”. **(Institutional Stakeholder in Kenya)**

On the other hand, CPs across the three countries were more interested in practical vaccine characteristics, stressing the need for clear and transparent information before adopting new vaccines:

> “*I won’t accept a new vaccine unless I know how it works. We need information on its advantages, side effects, mode and age for administration*”. **(FGD participant in Kenya)**

However, Ethiopian CPs demonstrated strong historical awareness of vaccine benefits, linking acceptance to past experiences with vaccine-preventable diseases:

> “*I don’t think there would be obstacles. The community knows well about benefits of vaccines from experience. The people have learned from past harms caused by diseases like smallpox and pertussis, so they have been taking their children for vaccine services*”. **(FGD participant in Ethiopia)**

##### Population-based evidence

The ISs stressed integrating social and cultural considerations in vaccine rollout strategies. They advocated for community engagement in vaccine policy decisions to enhance ownership and acceptance:

> “*We look at data around acceptability and even feasibility, particularly for GAVI, because we’ve been a GAVI-supported country for long. They are very strong and lay a lot of emphasis on what is called equity…. trying to make the vaccine reach as many children as possible where they need it…so again issues around equity*”. **(Institutional Stakeholder in Kenya)**

Interestingly, one ISs highlighted concerns about population estimate accuracy, noting that inconsistent denominators for vaccine needs and coverage can distort demand projections (e.g., over- or underestimation) and misrepresent immunisation performance:

> “*The other thing is improving population estimates, especially to calculate targets and vaccination coverage. Different denominators are used when calculating vaccine needs versus vaccine coverage making it difficult to determine accurate targets. Which one do you trust? As researchers, we need to support the ministry in addressing this*.” **(Institutional Stakeholder in Malawi)**

##### Programme readiness and logistics

The ISs in each country highlighted the need to assess the EPI delivery feasibility including funding, supply chains, cold chain capacity, health worker training, and surveillance systems to ensure successful vaccine integration:

> “*As a country we are highly donor-dependent so you must consider other critical factors. We need to establish whether we have adequately trained health workers and sufficient infrastructure. When we recommend any vaccine, we need to evaluate if the necessary systems and resources are in place to initiate and sustain this initiative*” **(Institutional Stakeholder in Malawi).**

Four Kenyan ISs questioned financial sustainability during GAVI transition, stressing EPI readiness assessments before new vaccine introduction:

> “*So, for a long time, we didn’t care too much because we are part of GAVI, but we are heading towards graduation from GAVI….. now it means at some point in the very near future, the country would have to meet the cost of those vaccines in addition to other existing vaccines that we’ve already introduced*.” **(Institutional Stakeholder in Kenya)**

#### 3. Trusted sources of evidence

The ISs universally recognised MoHs as trusted stewards of national health-related data, given their systematic collection through government health services:

> *So as a standard, all the data concerning health is owned by the government. So, data needs to come from the government…any data that you want to use for policy decisions should be data that is hosted by the government. So, because this data is collected usually, through routine health services and these are run by Ministry of Health, and they are sole owners of this data*. **(Institutional Stakeholder in Malawi)**

This was reinforced by an IS in Kenya who also emphasised the crucial role of research institutions in supporting the government in evidence generation for policy decisions:

> *So, most of the data is often collected from the health information system…the trusted institution is often government but also there are entities that have been known as research institutions that have conducted substantial amount of research over years and therefore there is an element of credibility*. **(Institutional Stakeholder in Kenya)**

Research institutions, such as the Kenya Medical Research Institute (KEMRI) in Kenya and the Malawi-Liverpool-Wellcome Trust (MLW) in Malawi, were recognised for producing high-quality data. When such data are published in peer-reviewed journals, they further enhance confidence in their credibility. Teaching and training institutions, including Addis Ababa University (AAU) in Ethiopia, was also cited as contributing valuable evidence, which could be used to support inform policies.

Additionally, partner institutions, including UN organisations were identified as trusted sources of data. This perspective is illustrated in Figure 3 and echoed below:

> “*Data should also come from the UN agencies WHO and UNICEF. Those are the other two organisations that are trusted to be able to provide that kind of data. I think the others would come from probably institutions that support vaccination, like GAVI, USAID and BMGF*.” **(Institutional Stakeholder in Ethiopia)**

**Figure 3:**
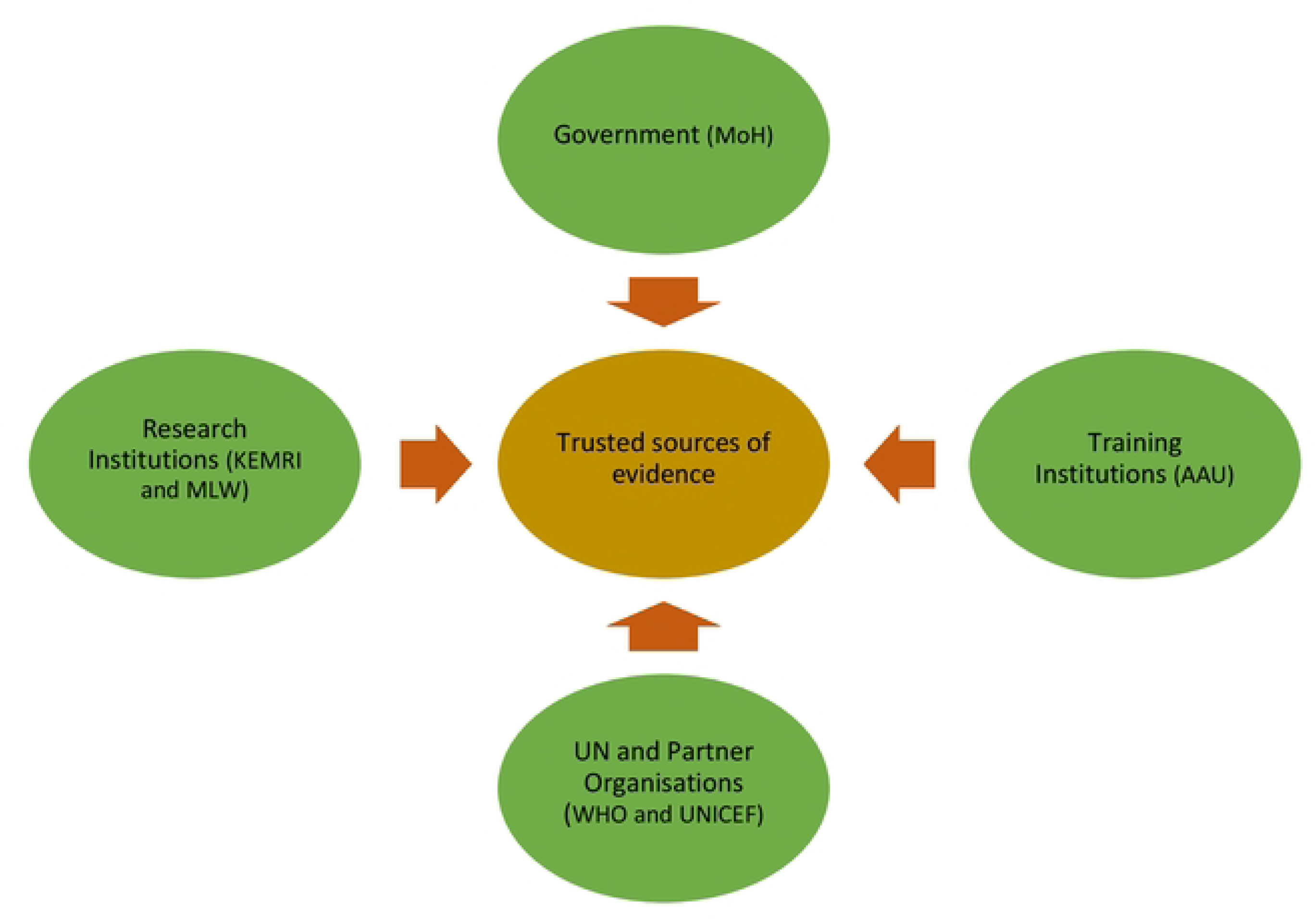
**Trusted sources of evidence for policy decisions**

#### 4. Strategies for increasing evidence use

ISs retaliated that evidence alone is insufficient and deliberate strategies are needed to integrate research into policymaking. Key approaches included:

##### Timely provision of evidence aligned with country needs

The ISs emphasised the need for timely, policy-stage-specific evidence (from agenda-setting to evaluation) highlighting the importance of aligning research priorities with decision-makers’ needs and ensuring accessibility:

> “*So, from the start, data must be available and adequate. Data is needed at different stages and there is different type of data that need to be provided at pre-policy formation stage, another during the implementation, monitoring, etc. So, I would say at all those time points, data is required*”. (**Institutional Stakeholder in Malawi)**

##### Engaging policymakers in research processes and early involvement of NITAGs

MoH ISs emphasised that engaging policymakers early in research builds trust and promotes evidence uptake. Similarly, NITAG ISs indicated that their involvement ahead of vaccine introductions allows comprehensive review of evidence and strengthen policy decisions:

> “*So, we are also becoming part of that research, for example, so that we can understand the whole process, which builds trust and helps the investigator understand the context. So, most of the time, from my experience, I can say that the evidence produced with the engagement of policymakers and programme people, like me and my expertise, is translated to policy*”. (**Institutional Stakeholder in Ethiopia)**

> *“What I would suggest the involvement of NITAG from the beginning, would be key If any new vaccine is to be introduced. If you get NITAG on board, to start discussing these issues about a particular pathogen or the potential for a vaccine, then they help you out to be the advocates and they start looking at evidence for introducing the vaccine early on”.* (**Institutional Stakeholder in Malawi**)

##### Strengthening National Public Health Institutes (NPHIs)

The ISs identified timeliness, quality, and accessibility challenges with routine national surveillance data and suggested the strengthening NPHIs as a key strategy to address these gaps:

> “*When it comes to data, some of the challenges that we are facing are issues with the quality of the data completeness, the timeliness of reporting, which we are working on to improve, but I think we still have opportunities for improvement as we move along*”. **(Institutional Stakeholder in Malawi)**

> “*Something which has been on the table for many years in Kenya and I think it’s the issues of the National Public Health Institute. And I think if established, it will be quite critical in terms of coordinating all these and to make sure that quality data is available for decision makers and that can be relied on when discussions on policy changes and directions need to be done*”. **(Institutional Stakeholder in Kenya)**

##### Prioritising diarrhoeal diseases and establishing technical working groups (TWGs)

The ISs emphasized prioritizing diarrhoeal diseases alongside malaria, tuberculosis, and HIV to ensure equitable public health focus and funding. They also recommended establishing diarrhoeal disease-specific TWGs within MoHs to strengthen evidence-based decision-making, complementing NITAGs by providing specialised platforms for in-depth review of vaccine-preventable diarrhoeal infections:

> “*The disease must be recognised as a major public health problem. If it is no more public health problem then it is very difficult to justify, for example, that’s what we have as a problem with regards to yellow fever in the country, because the government thinks that it is not a problem*”.

> (**Institutional Stakeholder in Ethiopia**)

> *“I think having something like a technical working group or coordinating unit just like other major infections like HIV and TB, I think it will help in sort of pushing and making sure that whatever is being done is being translated into actions that can help in alleviating gastrointestinal infections”*.

> (**Institutional Stakeholder in Malawi**)

##### Improving communication for knowledge mobilisation

The ISs emphasised that effective communication among policymakers, researchers, and the public is critical for evidence-based policy translation. They recommended tailored strategies, based on audience and information type, include face-to-face engagements, digital platforms (e.g., social media), policy briefs, and collaborative forums like advisory committees:

> “*There are multiple channels for communicating with stakeholders during the process of making policies, but what would define the best strategy would be segregating the target stakeholders that are being earmarked for influence and then the most appropriate channel would be defined based on that*”. **(Institutional Stakeholder in Kenya)**

Both ISs and CPs acknowledged that public engagement is crucial, noting that intermediaries such as journalists can enhance message dissemination. While CPs in all countries identified a need for education on new vaccines after highlighting knowledge gaps, with Malawian CPs recognised rotavirus and cholera vaccines, whereas some Kenyan and Ethiopian CPs were unaware their children had received rotavirus vaccination:

> “*Apart from administering the rota vaccine, there is also a policy that stipulates that all children under five years of age should be given a vaccine that protects them from cholera so that they cannot suffer from cholera. I would say that another vaccine for children under five years of age is the cholera vaccine*.” **(FGD participant in Malawi)**

> “*I don’t know the vaccines given to the child; I always give the doctor the child’s card, and the kid gets the vaccine*.” **(FGD participant in Kenya)**

> “*I just recognised the rota vaccine when you told us it is given in the form of droplets… The community doesn’t know the purpose of the vaccine and doesn’t know the side effects, and that is because the health workers don’t explain the side effects*.” **(FGD participant in Ethiopia)**

Despite these gaps, CPs in all three countries reported that health workers and community organisations are the primary sources of vaccine information. However, they emphasised the need for widespread campaigns involving trusted local leaders, such as chiefs and religious figures, when introducing new vaccines to foster trust and acceptance:

> “*I feel like if other vaccines are introduced, people would be happy But I would prefer that before any vaccine is introduced, people in the communities should be civic educated about how these vaccines work……the messages can be disseminated using religious leaders because most of the people trust their religious leaders most*”. **(FGD participant in Malawi)**

CPs recommended dissemination strategies including community gatherings, maternal training, radio campaigns, door-to-door outreach, and tailored advocacy. In Ethiopia, CPs emphasised targeted follow-ups and advocacy for vulnerable groups (e.g., street children):

> “*I have observed that the orphans living on the street don’t know what vaccines mean. The youths living there give birth to one on another, and no one follows up on them. We need to support and advocate for them*”. **(FGD participant in Ethiopia)**

## Discussion

This study explored institutional stakeholders’ and community perspectives on evidence needs and involvement in policy decisions for potential *Shigella* and ETEC vaccine introduction in Ethiopia, Kenya, and Malawi. Vaccine adoption is complex, shaped by political dynamics (36) but requiring robust evidence on disease burden, vaccine profiles, population-specific factors, and implementation feasibility. Key gaps include limited diarrhoeal disease-focused technical working groups and opportunities to strengthen evidence use and community engagement for inclusive policymaking. Disease burden data are critical for guiding intervention strategies, including vaccine adoption, while evidence on vaccine efficacy, safety, and cost-effectiveness is equally vital to justify prioritisation. Early availability of both datasets is key to accelerating policy decisions and enabling timely action, a principle well-supported by literature on vaccine introduction in LMICs (37, 38).

This study underscores the critical role of population-based evidence in guiding new vaccine introductions. Community attitudes, knowledge, and practices significantly impact vaccine acceptance, particularly in an era of widespread misinformation (39). Accurate, context-specific data is essential for developing effective communication strategies and fostering informed community engagement. Additionally, programmatic readiness, including staff capacity, cold chain functionality, and robust surveillance, also determines successful vaccine rollout. Existing immunisation coverage reflects a program’s capacity to integrate new vaccines (40), but accurate metrics depend on context-appropriate denominators. Standardising data sources and improving accuracy are vital for optimising resource allocation and program impact (41, 42). Furthermore, programmatic readiness hinges on a country’s political commitment to vaccine adoption and sustainability (43). For non-GAVI-supported countries, securing independent funding and delivery systems is critical. South Africa, the first African nation to introduce rotavirus and pneumococcal conjugate vaccines, demonstrated how such challenges can be overcome independently (44).

Our findings support La Torre *et al.’s* (45) recommendation for Health Technology Assessment (HTA) in vaccine introduction, which aligns with WHO and Burchett frameworks in emphasising multidisciplinary evidence (46, 47). We further validate Donadel *et al*. (37) on the critical need to consider programmatic factors, particularly in LMICs where resource allocation determines implementation success. This addresses a historical gap in decision-making frameworks that often neglected programmatic readiness despite its vital role in vaccine rollout (24).

Robust local evidence from credible research institutions to inform vaccine policy is critical as highlighted in our study. Yet the absence of local evidence often delays or prevents vaccine introduction, as seen in Indonesia’s rejection of the H1N1 vaccine due to limited local data, despite WHO-documented global burden(48, 49). To overcome such barriers, LMICs should strengthen research capacity by enhancing surveillance systems, updating national research agendas, and securing dedicated research funding for multidisciplinary studies.

Diarrhoeal diseases are among the top five causes of morbidity and mortality in sub-Saharan Africa, exceeding HIV and tuberculosis in Disability-Adjusted Life Years (DALYs) (50). Given their significant health burden, this study recommends prioritising them in public health by establishing diarrhoeal disease-specific technical working groups within Ministries of Health. Furthermore, integrating community involvement in these efforts can enhance advocacy for incorporating new vaccines into national immunisation programmes.

Our FGDs with CEI groups highlighted the need for clear, detailed vaccine information covering mechanisms, safety, side effects, schedules, and administration. Effective dissemination through trusted sources such as health workers and community leaders (e.g., chiefs, religious figures) was key to fostering parental trust aligning with findings from Ames *et al*. (51) and Oku *et al.* (52).

Additionally, meaningful community engagement in policy processes can enhance ownership, yet contextual reviews are needed to evaluate its actual impact. For example, a Malawi study found that while community involvement is often mandated, stakeholders viewed it as tokenistic, with their input rarely influencing decisions due to power imbalances (53). Similar evidence across Africa support the need to address barriers and leverage facilitators for effective participation (54).

### Future research

By creating awareness and demand, this work supports the development of potential *Shigella* and ETEC vaccines, which are currently in progress. Although the timeline for licensed vaccines remains uncertain, several candidates are advancing through preclinical and Phase 2 trials—including a recently concluded ETEC trial in The Gambia and an anticipated *Shigella* trial completion in 2025(55–57). Regulatory approvals and other factors will influence their availability, but the current progress is promising for near-future deployment.

Key research priorities include generating local *Shigella* and ETEC burden data (e.g., through the Enterics for Global Health *Shigella* surveillance study(58)), evaluating vaccine cost-effectiveness, and exploring community perspectives on diarrheal management and vaccine acceptance. Programmatic readiness, especially in Kenya’s post-GAVI transition, also warrants attention. Additionally, studies on community engagement in vaccine policy development are needed to strengthen ownership, acceptance, and alignment of new vaccine initiatives with community needs.

### Strengths and Limitations

This study employed a rigorous qualitative approach, using FGDs and IDIs to generate in-depth insights from institutional stakeholders and community representatives across three countries. These methods enabled a nuanced understanding of evidence needs for policymaking and the dynamics of community engagement in health policy processes. By capturing diverse stakeholder voices, the study highlights key factors influencing vaccine policy development and implementation in LMICs.

Several limitations should be noted. First, the purposive sampling strategy, while suitable for the study’s aims, limits generalisability and may have excluded other relevant perspectives. Second, conducting only one FGD per country constrained the breadth of community viewpoints; additional FGDs with varied community or civil society groups could have enriched the findings. Third, language translation and transcription may have led to the loss of subtle meanings or cultural nuances, despite measures to preserve the accuracy of participants’ responses.

Nevertheless, the study offers valuable insights into the evidence and engagement processes necessary to support *Shigella a*nd ETEC vaccine introduction. Future research should expand the range of community perspectives and stakeholder groups to strengthen and build on these findings.

## Conclusion

The introduction of new vaccines into national immunisation programs is a complex process influenced by various factors, with technical decisions requiring robust evidence. Institutional stakeholders emphasised the need for local, high-quality data on disease burden, vaccine characteristics, and programmatic readiness to support timely and informed policy decisions. Community participants generally supported vaccines, especially when tangible health benefits are observed. However, providing clear information about how new vaccines work, including their safety and efficacy, is crucial for broad acceptance.

Both ISs and CPs highlighted the importance of community engagement in policy development. However, such engagement remains limited in LMICs, often restricted to implementation. Therefore, meaningful community involvement in policy formulation is required, emphasising that such engagement could enhance ownership, trust, and compliance with new vaccine policies. This engagement is essential for achieving equitable and sustainable vaccine introduction.

### Policy and practice implications

- Governments and partners should prioritise investments in generating localised evidence for vaccine policy decisions.
- Mechanisms to institutionalise meaningful community involvement in national policy processes must be developed to foster trust and ownership.
- Sustained efforts to improve vaccine communication strategies are essential, with a focus on addressing misconceptions and leveraging trusted messengers.
- Countries should assess and strengthen their programmatic readiness, particularly considering financial sustainability challenges for new vaccines.
- TWGs for diarrhoeal diseases should be established in MoHs, with community representatives.

## Additional information

### Disclosure of interest statement

All authors declare no conflict of interest.

#### GHRG-GI consortium

Daniel Asrat (Addis Ababa University, Addis Ababa, Ethiopia); Prisca Benedicto (Malawi Liverpool Wellcome Programme, Blantyre, Malawi); Catherine Beavis (University of Liverpool, UK); Christina Bronowski (University of Liverpool, Liverpool, UK); Jobiba Chinkhumba (Kamuzu University of Health Sciences, Blantyre, Malawi); Helen Clough (University of Liverpool, Liverpool, UK); Jen Cornick (University of Liverpool, Liverpool, UK and Malawi Liverpool Wellcome Programme, Blantyre, Malawi); Khuzwayo Jere (University of Liverpool, Liverpool, UK); James Ngumo Karis (Kenya Medical Research Institute, Nairobi, Kenya); Sam Kariuki (Kenya Medical Research Institute, Nairobi, Kenya); Cecilia Mbae (Kenya Medical Research Institute, Nairobi, Kenya); Amha Mekasha (Addis Ababa University, Addis Ababa, Ethiopia); Chisomo Msefula (Kamuzu University of Health Sciences, Blantyre, Malawi); Edson Mwinjiwa (Malawi Liverpool Wellcome Programme, Blantyre, Malawi), Latif Ndeketa (University of Liverpool, Liverpool, UK and Malawi Liverpool Wellcome Programme, Blantyre, Malawi); Phelgona Otieno (Kenya Medical Research Institute, Nairobi, Kenya); Virginia Pitzer (Yale, New Haven, USA); Yemisrach Shumeye (Addis Ababa University, Addis Ababa, Ethiopia); Abebe Habtamu Tamire (Addis Ababa University, Addis Ababa, Ethiopia); Fred Were (Kenya Medical Research Institute, Nairobi, Kenya); Mengistu Yilma (Addis Ababa University, Addis Ababa, Ethiopia).

## Data Availability

All data requests should be submitted to the corresponding author for consideration. Access to anonymised data may be granted following review.

## Acknowledgement

We would like to express our sincere gratitude to all the participants in this study for their time and valuable contributions to the discussions.

## Patient data statement (if required)

This study did not utilise any patient data.

## Ethics statement

Ethical approvals for the study were obtained from the respective ethics review boards in each country: Department of Pediatrics at Addis Ababa University (030-23-ped) in Ethiopia, Kenya Medical Research Institute’s Scientific Ethical Review Unit (SERU NO: KEMRI/SERU/CMR/P00221-010-2022/4637) in Kenya, and the Kamuzu University of Health Sciences (P.10/22/3790) in Malawi. Additionally, ethical approval was granted by the University of Liverpool, UK (Reference number 12443).

Before participation in the IDIs and FGDs, participants were provided with detailed information about the study, given the opportunity to ask questions, and individual informed consent was obtained. Participants were also informed of their right to withdraw from the study at any stage without consequences.

## Information governance statement

A data governance plan was developed for this study to outline the research context and establish specific processes for ensuring effective and secure data management. The approach was structured around five key principles: safe projects, safe people, safe settings, safe data, and safe outputs.

At each level, potential data governance risks were identified, and appropriate controls were specified to mitigate risks and uphold data integrity, confidentiality, and ethical standards throughout the study.

## List of abbreviations

AMR: antimicrobial resistance
CEI: community engagement and involvement
CPs: community participants
ETEC: Enterotoxigenic *Escherichia coli*
EPIs: expanded programmes of immunisation
FGDs: focus group discussions
GHRG-GIs: Global Health Research Group on Gastrointestinal Infections
IDIs: in-depth interviews
ISs: institutional stakeholders
LMICs: low- and middle-income countries
MoH: ministries of health
NITAGs: National Immunization Technical Advisory Groups
NIHR: National Institute for Health and Care Research
RVV: rotavirus vaccine
SSA: sub-Saharan Africa
WASH: water, sanitation and hygiene

